# Epigenetic Aging Clocks Associate with Cognitive Status but Not Cognitive Decline: Evidence from the Parkinson’s Progression Markers Initiative

**DOI:** 10.64898/2026.05.11.26352912

**Authors:** Bowen Feng, Andy Gao, Jingyun Yang

## Abstract

Cognitive impairment is a major source of disability in Parkinsonian disorders, yet biomarkers that distinguish cognitive status from cognitive decline remain limited. DNA methylation-based epigenetic aging measures capture complementary dimensions of biological aging, but it remains unclear whether they primarily reflect stable differences in cognitive vulnerability or longitudinal cognitive change. We examined associations between epigenetic aging measures and global cognition in the Parkinson’s Progression Markers Initiative (PPMI) cohort. Seven epigenetic aging measures were derived from peripheral blood DNA methylation data, and cognition was assessed longitudinally using the Montreal Cognitive Assessment (MoCA). Linear mixed-effects models were applied in complementary frameworks, including baseline-plus-change-from-baseline models and within-person versus between-person decomposition models. Secondary analyses included baseline clock-by-time interaction models and a decline-focused sensitivity analysis. Across analyses, higher epigenetic aging was consistently associated with lower overall MoCA scores. In the baseline-plus-change-from-baseline models, the analytic baseline component showed the dominant signal, whereas the change-from-baseline terms were not significant after false discovery rate correction. In the within-person versus between-person decomposition models, associations were concentrated in the between-person component, while within-person deviation terms were not significant. Secondary analyses were consistent with this pattern. Together, these findings suggest that blood-based epigenetic aging measures may be more informative as biomarkers of cognitive status or vulnerability than as markers of short-term cognitive progression. Larger studies with longer follow-up and more detailed cognitive phenotyping are needed to clarify their longitudinal relevance.

## Introduction

Cognitive impairment is a major non-motor manifestation of Parkinsonian disorders and a key determinant of disability, loss of independence, reduced quality of life, and caregiver burden [1, 2]. However, cognitive trajectories are highly heterogeneous across individuals, spanning relatively stable function in some patients and progressive decline in others [1, 3]. This heterogeneity complicates prognostication and underscores the need for biomarkers that can characterize cognitive vulnerability, refine phenotypic stratification, and provide insight into underlying mechanisms [1, 4]. Biological aging is increasingly implicated in neurodegenerative processes, yet how systemic aging signatures relate to cognitive performance and cognitive change over time remains insufficiently understood [5, 6].

Among blood-based biomarkers of biological aging, DNA methylation-derived epigenetic clocks are of particular interest because they condense high-dimensional methylation information into quantitative measures that capture distinct dimensions of aging biology [7–13]. The first-generation clocks developed by Horvath and Hannum were primarily trained to predict chronological age and are often viewed as measures of age-related methylation patterns across tissues or, in the case of Hannum, in blood [7, 8]. Subsequently, second-generation measures were designed to move beyond chronological age prediction toward clinically relevant aspects of biological aging. PhenoAge incorporates information related to physiological dysregulation and morbidity risk, whereas GrimAge v1 and GrimAge v2 were developed to capture mortality- and healthspan-related risk using methylation surrogates of plasma proteins and smoking exposure [9–11]. In contrast, DunedinPACE was developed to estimate the pace of biological aging rather than cumulative age-related change, thereby reflecting the rate of aging rather than biological age per se [12]. More recently, deep-learning approaches such as AltumAge have sought to improve age prediction by modeling complex, non-linear relationships in methylation data [13]. Collectively, these measures represent overlapping but non-equivalent constructs, providing a useful framework for examining which epigenetic aging dimensions are most relevant to cognitive phenotypes.

A growing body of evidence suggests that epigenetic aging measures are relevant to cognitive health, but the nature of that relationship remains incompletely resolved. Across community-based and aging cohorts, accelerated epigenetic aging has been associated with poorer cognitive performance, greater risk of cognitive impairment or dementia, and, in some studies, steeper longitudinal decline, although the strength and consistency of these associations vary across cognitive domains, study populations, and clock types [14–17]. In particular, newer measures such as GrimAge-related clocks and DunedinPACE have often shown stronger associations with adverse neurocognitive outcomes than first-generation age-prediction clocks, suggesting that measures optimized for mortality risk, physiological dysregulation, or pace of aging may capture biology more closely related to cognitive vulnerability [14, 16, 17]. Nevertheless, important uncertainty remains. Much of the existing literature has been cross-sectional or has not clearly separated relatively stable between-person differences from within-person change over time [15, 18]. This distinction is critical for interpretation, because a biomarker associated primarily with between-person differences may be most useful for phenotypic stratification and risk characterization, whereas a biomarker that covaries with within-person change may be more informative for tracking progression [18].

In this study, we examined whether blood-based epigenetic aging measures are associated with cognitive performance and cognitive change over time in the Parkinson’s Progression Markers Initiative cohort (PPMI) [19]. Motivated by the limited understanding of whether epigenetic aging signals primarily reflect cognitive status or ongoing decline, we aimed to clarify the nature of these associations in a longitudinal setting. Specifically, we sought to determine whether epigenetic aging measures are more informative as markers of cross-sectional differences in cognitive vulnerability than as indicators of cognitive decline over follow-up [15, 18].

## Methods

### Data sources

All clinical and DNA methylation data used in this study were obtained from PPMI, an ongoing international, multicenter, longitudinal observational study established in 2010 to support biomarker discovery and characterization of disease progression in Parkinson’s disease and related at-risk states [19, 20]. PPMI was designed to generate deeply phenotyped longitudinal clinical and biospecimen data under a harmonized protocol with broad data sharing through a centralized repository.

To maximize available follow-up, we constructed the analytic dataset using a two-stage data extraction approach. Whole-blood DNA methylation data were derived from the PPMI Methylation Profiling dataset, including Projects 120 and 140, using the June 7, 2021 release. Corresponding phenotype, cognitive, and other clinical covariate data for participants with methylation data were then downloaded separately from the PPMI repository using records available through January 2026, thereby capturing longitudinal follow-up from study entry through that time point. The present analysis was based on the merged dataset generated from these two sources.

PPMI enrolls participants across several protocol-defined groups, including individuals with Parkinson’s disease, healthy controls, and prodromal or genetically enriched participants, and follows them with standardized clinical evaluations, imaging, and biospecimen collection at regular intervals [19]. This design provides a robust framework for investigating longitudinal heterogeneity in clinical phenotypes and biomarker profiles. The analytic framework is illustrated in **Figure 1**.

**Figure 1.**
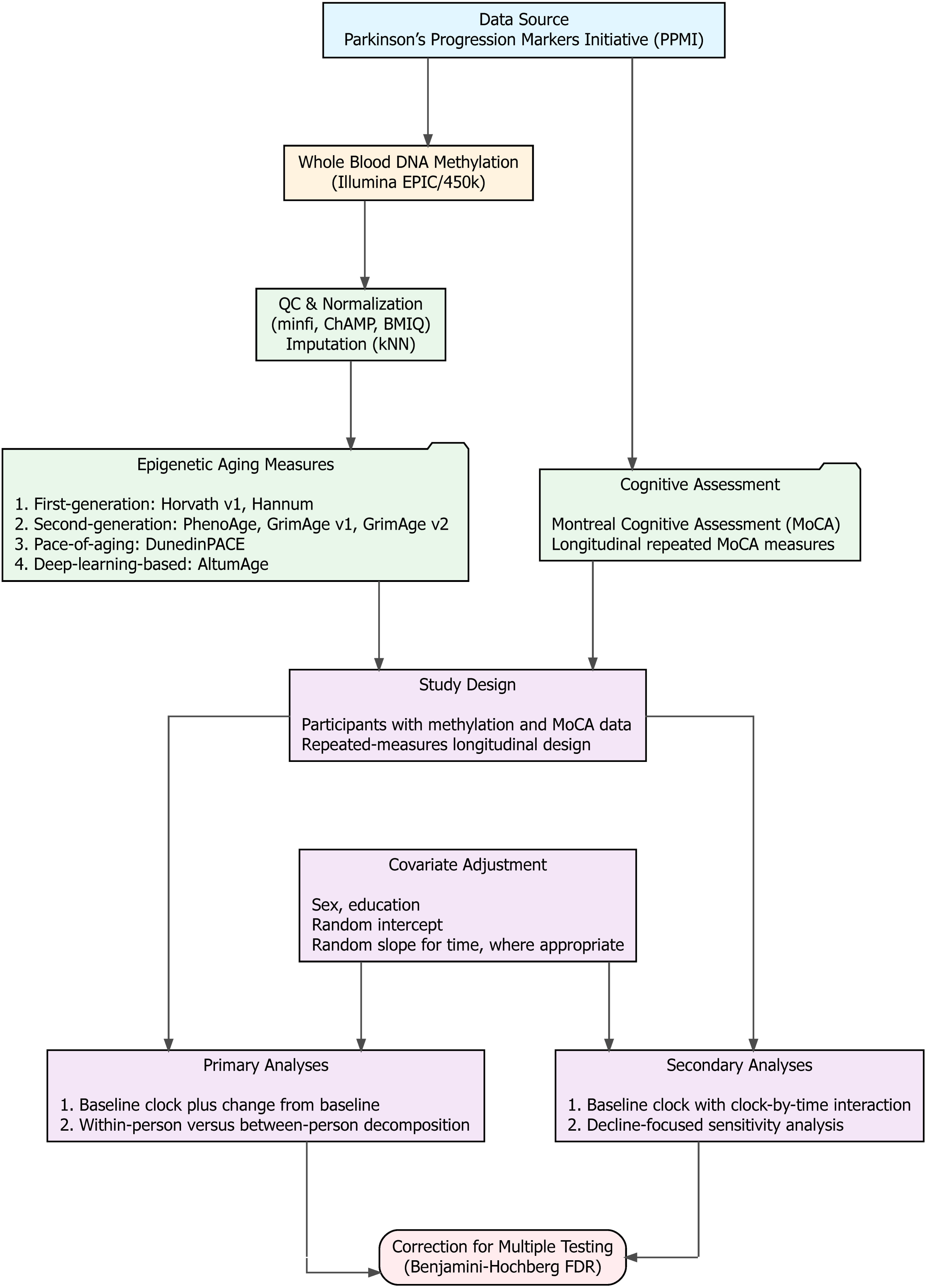
Overview of the study design and analytic framework. Longitudinal clinical assessments and whole-blood DNA methylation data were obtained from the PPMI. DNA methylation data underwent QC, normalization, imputation, and batch adjustment before the derivation of seven epigenetic aging measures. Cognitive performance was evaluated using repeated MoCA total scores. The analytic framework focused on participants with both epigenetic aging and MoCA data and used complementary longitudinal models to distinguish associations with cognitive status from associations with cognitive change. BMIQ, Beta-Mixture Quantile normalization; ChAMP, Chip Analysis Methylation Pipeline; FDR, false discovery rate; kNN, k-nearest neighbors imputation; MoCA, Montreal Cognitive Assessment; PPMI, Parkinson’s Progression Markers Initiative; QC, quality control.

### Assessment of cognition

Global cognition was assessed using the Montreal Cognitive Assessment (MoCA), a brief cognitive screening instrument developed to detect mild cognitive impairment and subtle cognitive deficits that may not be captured well by less sensitive screening tools [21]. The MoCA is a 30-point measure composed of items spanning multiple cognitive domains, including attention, executive function, memory, language, visuospatial ability, abstraction, and orientation, and typically requires about 10 minutes to administer [21, 22]. In the present study, the primary cognitive outcome was the total MoCA score.

Within PPMI, the MoCA is included as part of the standardized cognitive assessment battery and has been widely used as a measure of global cognitive function in longitudinal studies of early Parkinson’s disease [23–25]. Previous work has shown that early cognitive deficits in Parkinson’s disease may be subtle and that the MoCA is more sensitive than the Mini-Mental State Examination for detecting mild cognitive impairment in this setting [21, 26]. In PPMI studies, MoCA has been used both to characterize baseline cognitive status and to track cognitive change over time, with repeated assessments obtained during longitudinal follow-up, generally on an annual basis [23–25].

### DNA methylation data

Peripheral whole-blood samples were collected in PPMI at baseline and subsequent protocol-defined follow-up visits. DNA methylation data were obtained from the PPMI Methylation Profiling dataset (Projects 120 and 140), which provides raw Illumina Infinium array .idat files. Most samples were assayed using the Illumina HumanMethylationEPIC BeadChip. Because repeated blood collection is embedded in the longitudinal design of PPMI, some participants contributed multiple DNA methylation assessments over time, enabling evaluation of within-person biological change.

Raw Illumina Infinium files were processed using the minfi and ChAMP pipelines [27, 28]. Samples with incomplete raw intensity files were excluded. Standard quality-control procedures were applied at both the sample and probe levels, followed by imputation for missing values. Beta values were normalized using BMIQ to reduce probe design bias [29], and batch effects were adjusted using ComBat before generation of the final beta-value matrices for downstream analysis [30]. Additional details of the DNA methylation preprocessing procedures are described in a previous study [31].

### Calculation of epigenetic clocks

Using post-processed DNA methylation data, we calculated seven epigenetic aging measures representing complementary dimensions of biological aging, including first-generation chronological age predictors (Horvath v1 and Hannum) [7, 8], second-generation morbidity- and mortality-related clocks (PhenoAge, GrimAge v1, and GrimAge v2) [9–11], a pace-of-aging measure (DunedinPACE), and a deep-learning-based estimator (AltumAge) [13].

Epigenetic clock estimation was performed primarily in R, with AltumAge implemented in Python. Calculations followed procedures recommended by the original developers, including standard calibration steps where applicable. Additional technical details on clock calculation procedures and the interpretation of epigenetic clocks have been described previously [31].

### Statistical analysis

Participant characteristics were summarized using standard descriptive statistics. Categorical variables were presented as counts and percentages, whereas continuous variables were summarized using means and standard deviations.

The primary outcome was longitudinal global cognitive performance, measured by the total MoCA score. For all association analyses, each epigenetic aging measure was examined separately in linear mixed-effects models to account for repeated observations within participants. For each clock-specific analysis, the analytic baseline was defined as the first visit at which both MoCA total score and the corresponding epigenetic aging measure were available. Follow-up time was calculated in years from this analytic baseline. Sex and years of education were included as fixed-effect covariates in all models. Random intercepts were used to account for between-subject differences in cognitive level, and random slopes for follow-up time were incorporated where appropriate to allow subject-specific variation in cognitive trajectories.

Our primary analyses were designed to distinguish associations with cognitive level from associations with cognitive change and to separate stable between-person differences from within-person variation in epigenetic aging measures. First, we fit models that simultaneously included the analytic baseline value of each epigenetic aging measure and its change from the analytic baseline at each follow-up visit. In these models, the baseline term captured differences in epigenetic aging at the analytic baseline, whereas the change-from-baseline term evaluated whether longitudinal deviations from the analytic-baseline epigenetic aging measure were associated with concurrent differences in MoCA. Second, we fit within-person versus between-person decomposition models in which each epigenetic aging measure was partitioned into a subject-specific mean across visits and a time-varying deviation from that mean. In this framework, the subject-specific mean represented the between-person component, whereas the deviation term represented the within-person component. Together, these two primary analyses were intended to clarify whether epigenetic aging measures were more strongly associated with cognitive level than with longitudinal cognitive change.

We also performed two complementary secondary analyses. In one set of models, analytic baseline epigenetic aging, follow-up time, and their interaction were included to evaluate whether analytic baseline aging measures were associated with both overall MoCA level and rate of cognitive change over time. In additional decline-focused sensitivity analyses, the interaction between analytic baseline epigenetic aging and follow-up time was examined, while additionally adjusting for change from the analytic baseline in the epigenetic aging measure, to evaluate whether analytic baseline epigenetic aging remained associated with cognitive decline after accounting for contemporaneous change in the clock. These secondary analyses were included to provide complementary perspectives on the relationship between epigenetic aging measures and longitudinal cognitive trajectories.

Multiple comparisons were adjusted using the false discovery rate (FDR). All statistical analyses were performed using R version 4.4.2 [32].

## Results

**Table 1** provides the basic characteristics of the study participants. At the analytic baseline, the analytic sample included 570 participants contributing 1,513 MoCA observations. The mean age was 64 ± 9 years, and 59% were female. The cohort was predominantly White (96%) and highly educated, with a mean of 16 ± 4 years of education. Global cognitive performance was generally preserved at study entry, with a mean MoCA total score of 27 ± 3. The mean follow-up duration was 1.7 ± 0.6 years. Baseline values of the seven epigenetic aging measures are also summarized in **Table 1**.

**Table 1.**
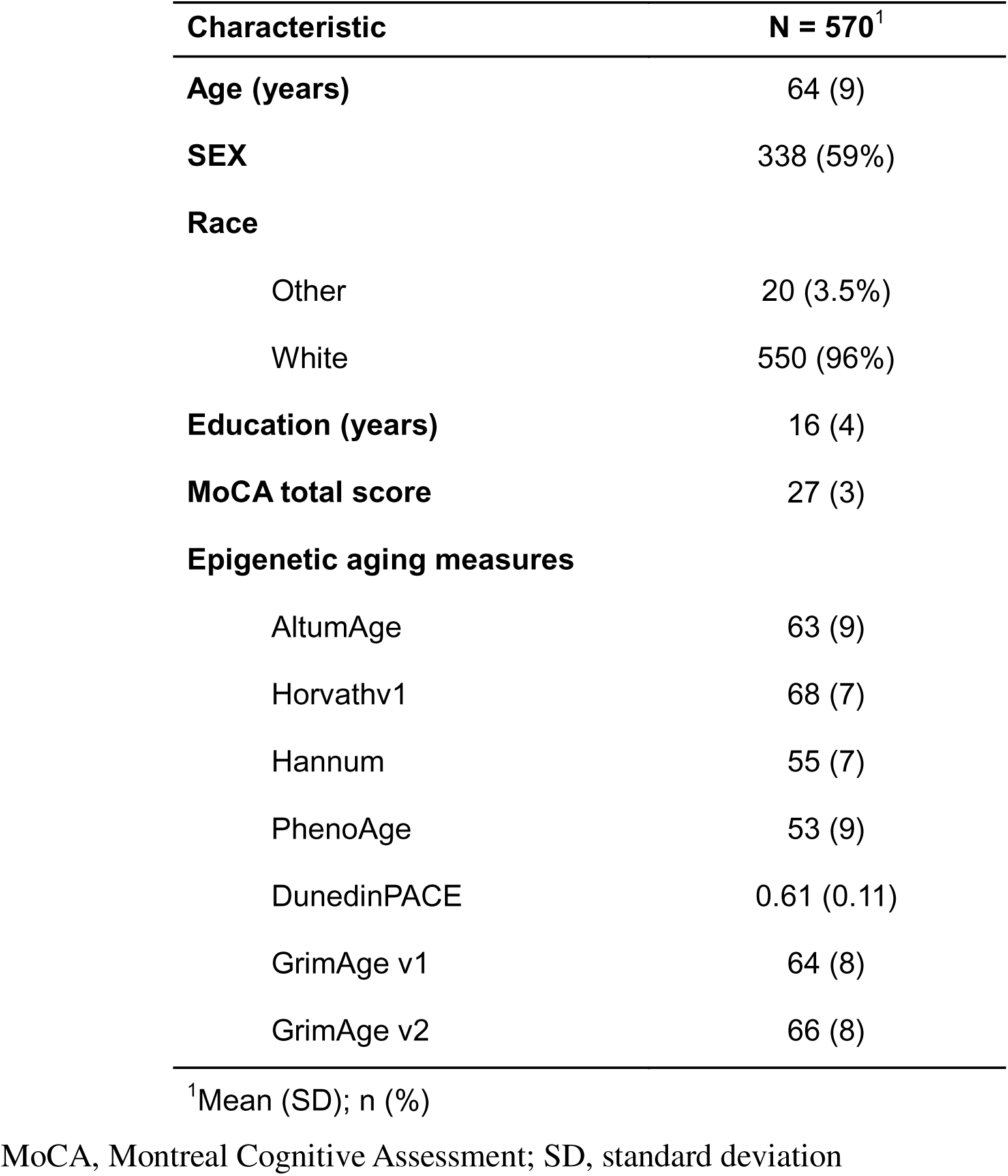
Baseline characteristics of the study participants.

### Primary analyses

In the primary baseline-plus-change-from-baseline models, the dominant signal was the analytic baseline component rather than the longitudinal change in the epigenetic aging measures. Analytic baseline epigenetic aging was negatively associated with MoCA for all seven clocks after FDR correction (**Figure 2**; **Table S1**). For most age-based clocks, coefficients ranged from −0.836 to −1.027, whereas DunedinPACE showed the same direction of association with a smaller coefficient on its native scale (β = −0.592, SE = 0.104, p = 1.736 × 10^-8^). In contrast, the change-from-baseline terms were not significant after FDR correction for any clock, although Horvath v1 showed a nominal association before correction (β = −0.072, SE = 0.028, p = 0.009, FDR-adjusted p = 0.065). Overall, these findings indicate that analytic-baseline differences in epigenetic aging were more strongly related to global cognitive level than were within-person changes relative to the analytic baseline.

**Figure 2.**
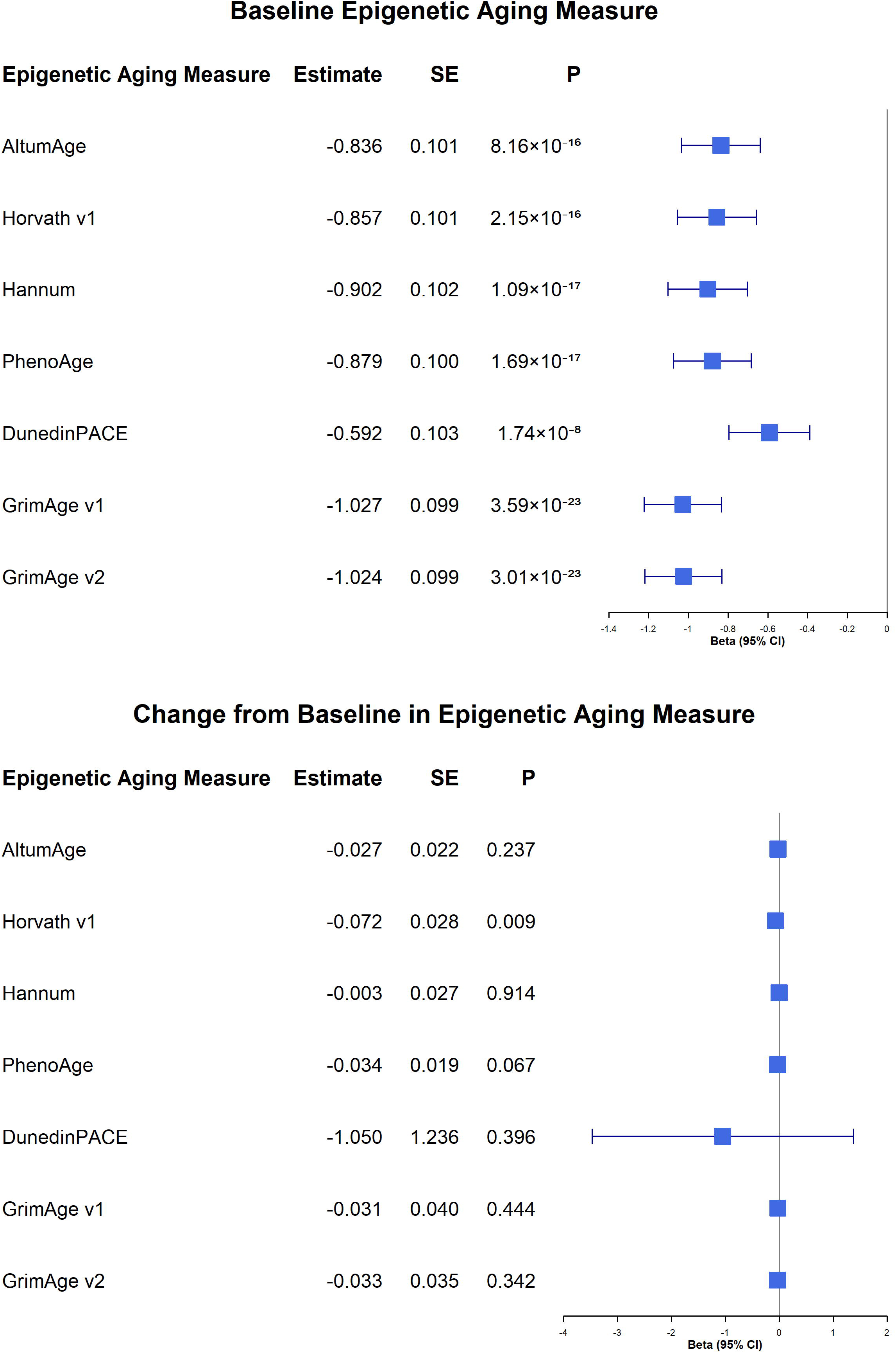
Baseline-plus-change-from-baseline associations between epigenetic aging measures and longitudinal MoCA scores. Forest plots show results from linear mixed-effects models evaluating analytic-baseline epigenetic aging measures and change from analytic baseline in relation to longitudinal MoCA total scores. The upper panel presents associations for analytic-baseline epigenetic aging measures, and the lower panel presents associations for change-from-baseline terms. MoCA, Montreal Cognitive Assessment; SE, standard error.

In the within-person versus between-person decomposition models, the between-person component showed a highly consistent association with cognitive level across all seven clocks, whereas the within-person component did not. For example, higher subject-specific mean AltumAge was associated with lower MoCA (β = −0.787, SE = 0.111, p = 3.562 × 10^-12^), and similar between-person associations were observed for the other epigenetic aging measures (**Figure 3**; **Table S2**). DunedinPACE showed the same direction of association, although its coefficient was smaller in magnitude because of its different scale (β = −0.616, SE = 0.118, p = 2.345 × 10^-7^). By contrast, the within-person deviation terms were not significant after FDR correction for any clock. For instance, the within-person AltumAge term was β = −0.341 (SE = 0.373, p = 0.361). These results indicate that the observed associations were predominantly between-person rather than within-person in nature (**Figure 3**; **Table S2**).

**Figure 3.**
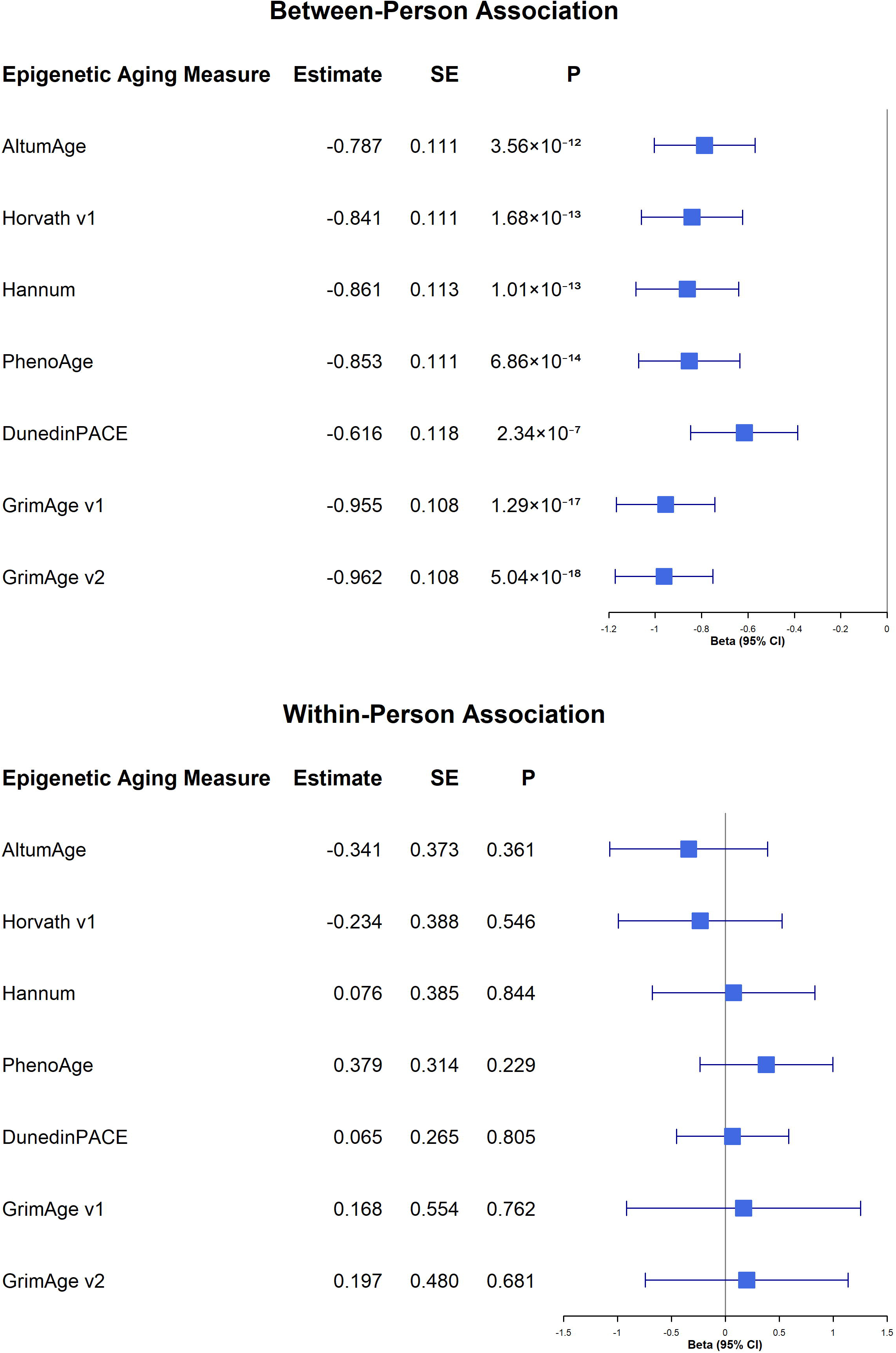
Within-person and between-person associations of epigenetic aging measures with longitudinal MoCA scores. Forest plots show results from linear mixed-effects models decomposing each epigenetic aging measure into between-person and within-person components. The upper panel presents the between-person component, defined as the subject-specific mean epigenetic aging measure across visits. The lower panel presents the within-person component, defined as the visit-specific deviation from the subject-specific mean. MoCA, Montreal Cognitive Assessment; SE, standard error.

### Secondary analyses

The secondary analyses were consistent with these primary results. In the analytic baseline clock-by-time interaction models, baseline clock main effects were again strongly negative and significant after FDR correction for all seven clocks. For most clocks, the effect sizes were in a broadly similar range (−0.801 to −0.975), whereas DunedinPACE showed a smaller coefficient magnitude on its native scale (β = −0.605, SE = 0.112, p = 1.008 × 10^-7^) (**Supplementary Figure 1**; **Table S3**). However, analytic baseline clock-by-time interaction terms did not survive FDR correction for any clock (**Supplementary Figure 1**; **Table S3**). These models, therefore, supported a robust association between analytic baseline epigenetic aging and overall cognitive level, but did not provide strong evidence that higher analytic baseline epigenetic aging was associated with the rate of MoCA decline.

A similar pattern was observed in the decline-focused sensitivity analysis (**Supplementary Figure 2**; **Table S4**). In that model, analytic-baseline clock effects remained significant after FDR correction for all seven epigenetic aging measures. For most age-based clocks, coefficients ranged from −0.803 to −0.976, whereas DunedinPACE again showed the same direction with a smaller coefficient on its native scale (β = −0.606, SE = 0.112, p = 9.588 × 10^-8^). The analytic-baseline clock-by-time interaction term was not significant for any clock, indicating limited evidence that higher analytic-baseline epigenetic aging was associated with faster MoCA decline. Change-from-baseline terms also did not survive FDR correction for any clock, although Horvath v1 showed a nominal association before correction (β = −0.074, SE = 0.028, p = 0.009, FDR-adjusted p = 0.063). Thus, even in a model emphasizing decline, the results continued to support associations with analytic-baseline or between-person differences in cognitive level, rather than with within-person clock change or cognitive decline over time.

## Discussion

In this longitudinal study of the PPMI cohort, we examined whether blood-based epigenetic aging measures were associated with global cognitive performance and cognitive change over time. Across complementary analytic frameworks, the most consistent finding was that higher epigenetic aging was associated with lower overall MoCA scores, whereas there was little evidence that within-person change in the clocks tracked cognitive decline. The within-person versus between-person decomposition further showed that these associations were driven predominantly by stable between-person differences rather than within-person variation over follow-up. Together, these findings suggest that epigenetic aging measures may be more informative as biomarkers of cognitive status or vulnerability than as markers of short-term cognitive progression. More broadly, the results indicate that systemic biological aging signals captured in blood may reflect inter-individual heterogeneity in cognitive function within PPMI.

A growing body of literature supports an association between epigenetic aging and cognitive health, but the pattern emerging across studies is not entirely uniform. In community-based and aging cohorts, accelerated epigenetic aging has been associated with poorer global cognition, greater risk of cognitive impairment or dementia, and, in some studies, steeper longitudinal cognitive decline [14–17]. These associations have often been stronger for second-generation clocks, such as GrimAge, and for pace-of-aging measures, such as DunedinPACE, than for first-generation chronological age predictors, suggesting that clocks optimized for morbidity, mortality, or aging rate may better capture the biology relevant to neurocognitive vulnerability [14, 16, 17]. Our findings are broadly consistent with this literature in that greater epigenetic aging was associated with worse cognitive performance. However, they extend prior work by showing that, within PPMI, the associations of epigenetic aging with cognition were more evident for overall cognitive level and between-person heterogeneity than for within-person change or cognitive decline over follow-up. This distinction may help explain some inconsistency in the prior literature, because many earlier studies did not explicitly separate between-person from within-person effects or compare baseline, change-from-baseline, and decomposition models within the same cohort [15, 18]. In this context, our study adds to the field by suggesting that epigenetic aging measures may be more informative as markers of cognitive status or vulnerability than as markers of short-term cognitive progression.

One reason our results may differ from studies reporting stronger associations with cognitive decline is that those studies were conducted in populations with different age structures, baseline clinical profiles, follow-up durations, and cognitive outcomes. In a previous twin study, participants were middle-aged men with a mean baseline age of 56 years and were followed for an average of 11.5 years. In that setting, Horvath-based age acceleration was associated with greater decline in executive and memory composite scores, despite no baseline cross-sectional association [15]. By contrast, Phyo et al. studied community-dwelling older adults aged 70 years or older and reported that GrimAge-related and pace-of-aging measures were associated with longitudinal deterioration in several domain-specific cognitive outcomes and incident dementia over 7 to 9 years of follow-up [16]. Compared with those cohorts, PPMI represents a distinct longitudinal research setting that includes participants with and without PD. The analytic sample in our study was relatively high functioning at study entry, with a mean baseline age of 64 years and a mean analytic baseline MoCA score of 27 [33, 34]. These differences are likely relevant because older cohorts with longer follow-up and domain-specific neuropsychological outcomes may be better positioned to detect gradual decline signals, whereas a relatively high-functioning cohort assessed primarily with a global screening measure such as MoCA may be more likely to reveal stable between-person differences in cognitive status than robust within-person decline effects over the observed interval. In addition, the prior studies largely related baseline methylation-based aging to subsequent cognitive change, whereas our study incorporated repeated methylation measures and explicitly separated baseline, change-from-baseline, between-person, and within-person components. Taken together, these design differences may help explain why prior studies identified stronger associations with cognitive decline, whereas our analyses more consistently highlighted associations with cognitive level and between-person heterogeneity.

Across our analyses, all seven epigenetic aging measures showed associations with cognitive status in the same direction, and for most clocks, the coefficient magnitudes were also broadly similar, with DunedinPACE showing the same directional pattern but a smaller coefficient on its native scale. This consistency is notable because these measures were developed using different training targets and modeling strategies, yet prior work suggests that they can still converge on related dimensions of organism-level aging.

PhenoAge, for example, was derived from a phenotypic age metric based on chronological age plus nine clinical biomarkers, thereby embedding signals of metabolic, inflammatory, hematologic, and physiological dysregulation [9]. GrimAge v1 was trained using a methylation-based smoking pack-years estimator together with methylation surrogates for plasma proteins, and GrimAge v2 further incorporated additional DNAm surrogates, including surrogates for C-reactive protein and hemoglobin A1c [10, 11].

DunedinPACE was developed to quantify the pace of biological aging from longitudinal change across 19 biomarkers spanning multiple organ systems and has also been associated with cognitive impairment and dementia risk [17]. Even among the earlier age-prediction clocks, comparative work suggests that Hannum’s clock is more closely tied to blood and immune-related aging processes, whereas Horvath’s clock captures broader pan-tissue aging signals [35]. Taken together, these findings suggest that the similar directional associations observed in our study may reflect the fact that, despite their methodological differences, these measures each capture related facets of systemic aging biology that are relevant to cognitive status.

Several limitations should be considered when interpreting these findings. Although the use of repeated methylation and cognitive assessments is a strength, the overall sample size remained modest for a longitudinal biomarker study, potentially reducing power to detect weaker clock-specific associations, particularly for within-person effects. The duration of follow-up may also have been insufficient to capture substantial within-person change in either epigenetic aging measures or MoCA performance. This issue is especially relevant in a relatively high-functioning cohort such as ours, in which analytic baseline MoCA scores were generally high, potentially limiting sensitivity to short-term decline and making stable between-person differences easier to detect than longitudinal change [19, 33, 36]. In addition, cognition was assessed using the MoCA total score, which provides a practical measure of global cognitive status but is less granular than a full neuropsychological battery. As a result, subtle domain-specific decline, particularly in executive, memory, or processing-speed functions, may not have been fully captured [37, 38].

Generalizability is also limited. PPMI is an intensively phenotyped research cohort that is predominantly White and highly educated, and these cohort characteristics may not reflect the broader population of individuals with Parkinson’s disease or related at-risk groups [19, 36, 39]. Social and structural determinants of health are also relevant, because epigenetic aging measures have been linked to socioeconomic adversity, education, and race/ethnicity in prior studies [40, 41]. All epigenetic aging measures in this study were derived from peripheral blood. Although blood-based clocks capture systemic aging processes relevant to health and disease, they do not necessarily reflect tissue-specific aging processes in the brain, and the correspondence between blood and brain for methylation-based aging measures is imperfect [42, 43]. Finally, as in all observational studies, residual confounding cannot be excluded. Our parsimonious models were designed to align with the conceptual structure of the epigenetic aging measures, but unmeasured factors, including lifestyle, comorbidity burden, medication exposure, and other disease-related characteristics, may still have influenced the observed associations. Because the analytic sample included both PD and non-PD participants, underlying heterogeneity across participant groups may also have attenuated disease-specific associations. Accordingly, the present findings should be interpreted as evidence of association rather than causation.

## Conclusions

In summary, blood-based epigenetic aging measures were consistently associated with global cognitive status in the PPMI cohort, whereas evidence for associations with cognitive decline over time was limited. These findings suggest that epigenetic aging measures may be more informative as biomarkers of cognitive vulnerability or status than as markers of short-term cognitive progression. Future studies with larger samples, longer follow-up, and more detailed domain-specific cognitive assessments are needed to clarify the longitudinal relevance of epigenetic aging and to determine whether these measures provide additional value for risk stratification and multimodal biomarker modeling in neurodegenerative research.

## Statements & Declarations

### Author Contributions

**Bowen Feng:** Formal analysis, Visualization, Writing – original draft

**Andy Gao:** Data curation, Visualization, Writing – original draft

**Jingyun Yang:** Conceptualization, Supervision, Writing – original draft, Writing – review and editing

### Ethics approval

This study used only publicly available data. As a result, ethical approval and consent to participate are not needed.

### Data Availability

The PPMI data generated or analyzed during this study are publicly available and can be requested at https://ida.loni.usc.edu/.

### Consent to participate

NA.

### Consent to publish

NA.

### Declaration of generative AI and AI-assisted technologies in the writing process

During the preparation of this work, the author(s) used ChatGPT (OpenAI)/Gemini to improve the clarity and readability of the language. After that, the author(s) carefully reviewed and edited the content to ensure accuracy and appropriateness and take full responsibility for the content of the published article.

---

**Supplementary Figure 1. Baseline epigenetic aging measures, clock-by-time interactions, and longitudinal MoCA scores.**

Forest plots show results from linear mixed-effects models evaluating analytic-baseline epigenetic aging measures and their interactions with follow-up time in relation to longitudinal MoCA total scores. The upper panel presents associations for analytic-baseline epigenetic aging measures, reflecting differences in overall MoCA level. The lower panel presents clock-by-time interaction terms, reflecting whether analytic-baseline epigenetic aging was associated with the rate of MoCA change over follow-up.

MoCA, Montreal Cognitive Assessment; SE, standard error.

**Supplementary Figure 2. Decline-focused sensitivity analysis of epigenetic aging measures and longitudinal MoCA scores.**

Forest plots show results from decline-focused sensitivity models evaluating the associations of analytic-baseline epigenetic aging measures with MoCA total scores, while additionally accounting for change from the analytic baseline in the corresponding epigenetic aging measure. The upper panel presents the main effects of analytic-baseline epigenetic aging measures, reflecting their associations with overall MoCA level. The lower panel presents the interaction terms between analytic-baseline epigenetic aging measures and follow-up time, reflecting whether higher analytic-baseline epigenetic aging was associated with the rate of MoCA change over time. Models were adjusted for sex and education, with random intercepts and random slopes for follow-up time where appropriate.

MoCA, Montreal Cognitive Assessment; SE, standard error.

