## Supplementary figures and images for "Epigenetic Aging Clocks Associate with Cognitive Status but Not Cognitive Decline: Evidence from the Parkinson’s Progression Markers Initiative"

### Supplementary Figure 1

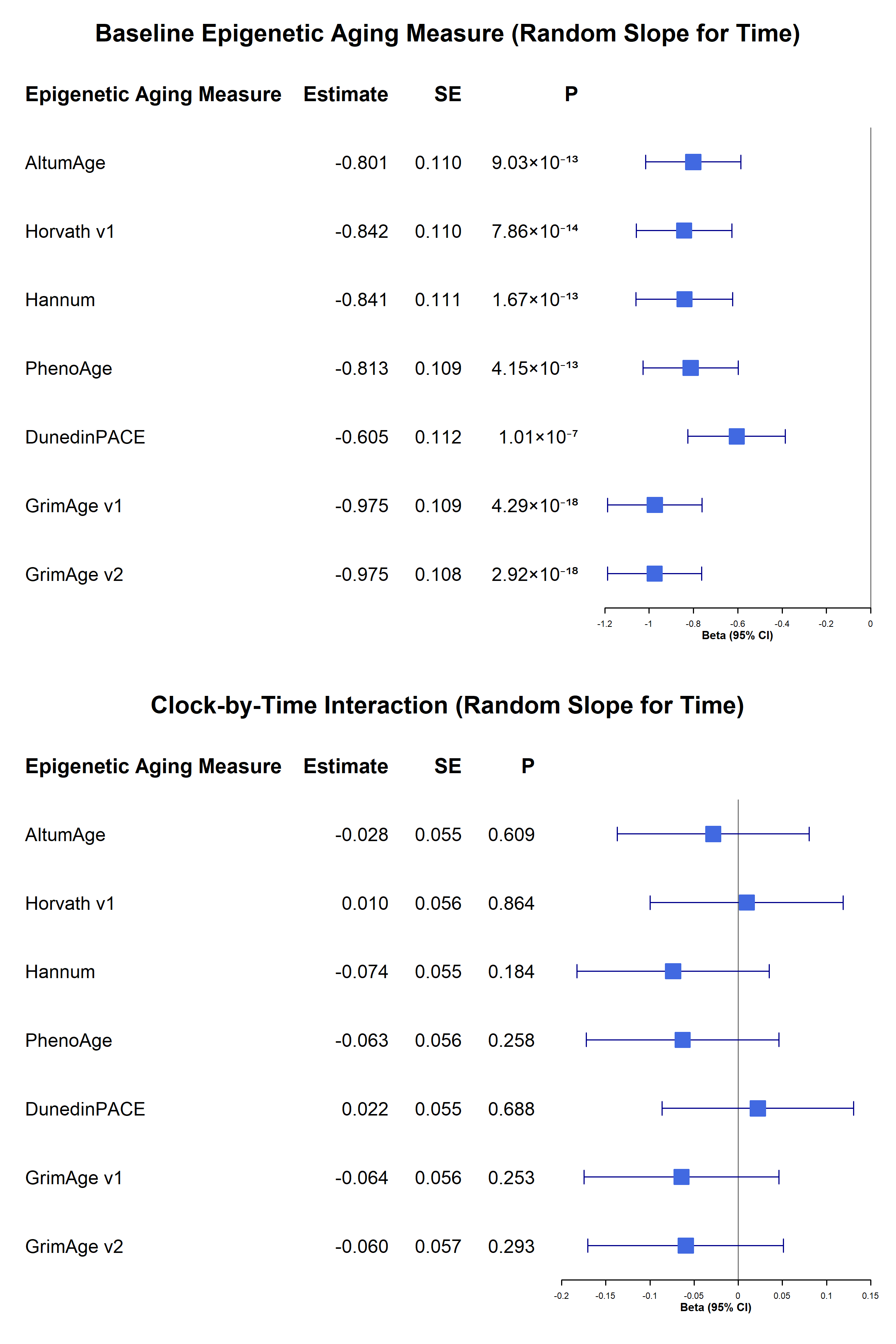

### Supplementary Figure 2

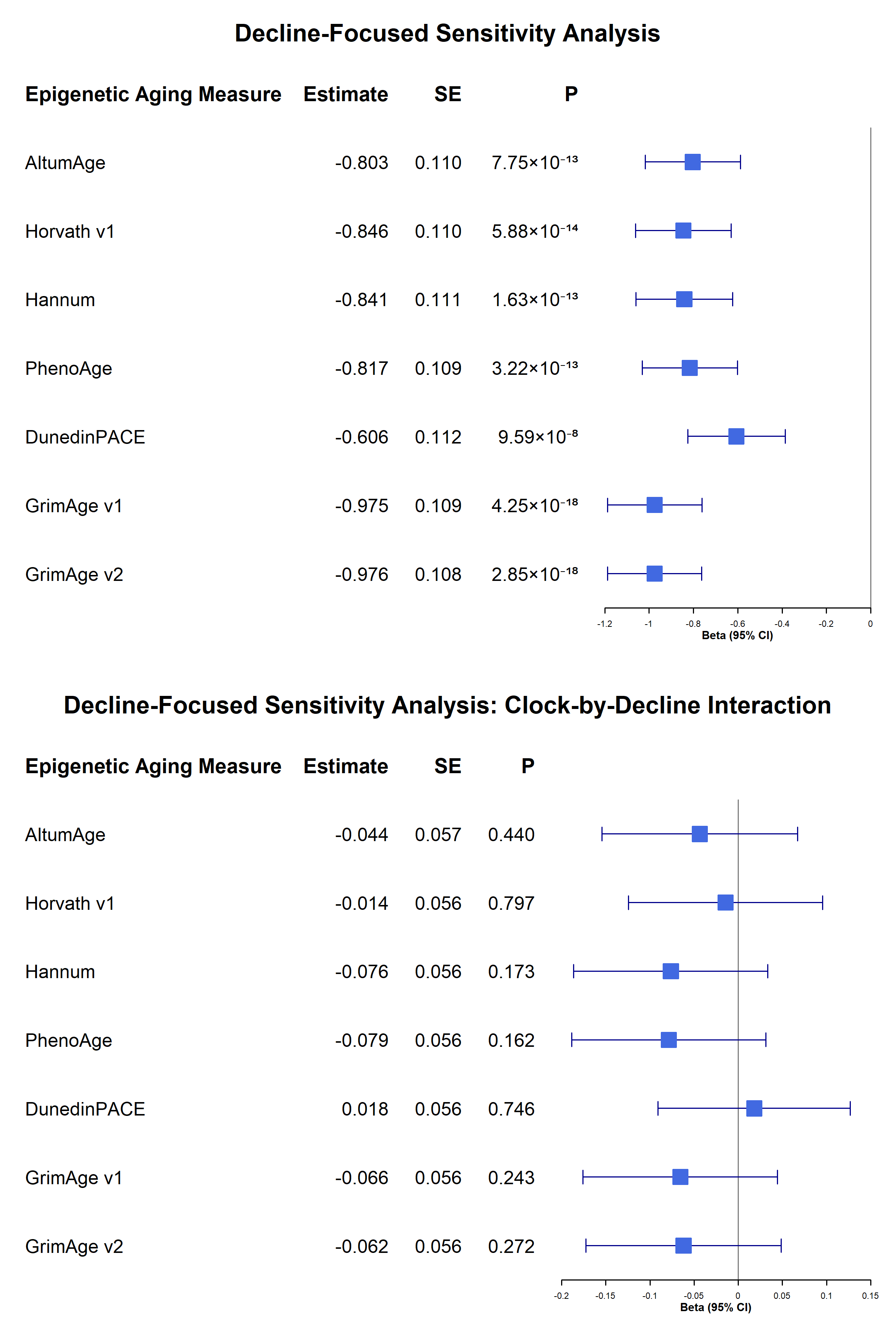
